# Outcomes from the Victorian Healthy Homes Program: a randomised control trial of home energy upgrades

**DOI:** 10.1101/2024.07.24.24310955

**Authors:** Katie Page, Lutfun Hossain, Dan Liu, Yohan Kim, Kerryn Wilmot, Patricia Kenny, Margaret Campbell, Toby Cumming, Scott Kelly, Thomas Longden, Kees van Gool, Rosalie Viney, VHHP team

**Affiliations:** Centre for Health Economics Research and Evaluation, University of Technology Sydney, Australia; Institute for Sustainable Futures, University of Technology Sydney, Australia; Sustainability Victoria, Melbourne, Australia

**Keywords:** housing, health, insulation, home upgrades (retrofits), randomised control trial, temperature, thermal comfort, Australia

## Abstract

**Introduction:** The Victorian Healthy Homes Program (VHHP) is the first randomised control trial (RCT) in Australia that investigates the impact of thermal home upgrades on energy and health outcomes in vulnerable individuals over winter in Victoria Australia.

**Methods:** A staggered parallel-group RCT design of 984 vulnerable households. The intervention group received their upgrade prior to their winter of recruitment and the control group received their upgrade after the winter of their recruitment.

**Results:** A relatively low-cost and simple home upgrade (average cost AU$2,809) resulted in reduced gas consumption (−25.5 MJ/day) and increased indoor winter temperatures (average daily increase of 0.33°C), and a reduction of exposure to cold conditions (<18°C) by an average of 0.71 hours (43 minutes) per day. The intervention group experienced improved mental health as measured by the SF-36 mental component summary and social care related quality of life measured by adult social care outcomes toolkit (ASCOT), and lower healthcare costs overall (an average of AU$887 per person) over the winter period.

**Conclusions:** The home upgrades significantly increased average winter indoor temperature, improved mental health and social care-related quality of life, and made householders more comfortable while bringing about reductions in overall healthcare use and costs.

**STRENGTHS AND LIMITATIONS OF THIS STUDY:**

- First randomised control trial of its kind in Australia to study the impact of thermal home upgrades on energy and health outcomes.
- The use of RCTs for such interventions is uncommon but provides a powerful approach to evaluation that can minimise the effects of confounding.
- The program was specifically designed to target vulnerable populations. These are groups with the most potential to benefit from home upgrades, either because of their socio-economic status or because of their chronic health conditions, or both.
- The retrofitted upgrade was tailored to each home based on need and delivered for a modest budget (up to AU$3,500)
- COVID-19 impacted the power of the study because not all upgrades were able to be delivered prior to winter. This was particularly relevant for the secondary health outcome measures.

## INTRODUCTION

Housing is a critical aspect of our lives and one indicator of the health and wealth of a population(1). Despite Australia’s relative wealth, the quality of its housing stock is very poor, with more than 91% of existing houses having an energy efficiency rating below code compliance(2). Measures to improve thermal comfort and residential energy efficiency can improve household indoor temperatures which in turn reduce the risk of adverse health outcomes(3).

There is growing recognition of the links between the home environment and health outcomes (3-8). International evidence suggests that there is an association between residential energy efficiency and negative morbidity and mortality outcomes, particularly during winter(9, 10). For example, a study from the UK study showed seasonal variation in mortality risk which differed by residential energy efficiency, with more energy efficient homes having lower mortality risk than homes with poor energy efficiency(10).

Health risks associated with cold indoor temperatures tend to have a greater impact on specific population groups including those with cardio-respiratory disease, children, and older people (8, 11). Indoor humidity levels have the potential to affect health adversely, for example through exacerbation of asthma and allergies. Vulnerable people, including the elderly, and those with a disability or chronic illnesses are at higher risk as they are more likely to spend most of their time at home and be more exposed to health risks associated with cold homes. Those with low incomes have limited means to improve the quality of their homes or afford increasing heating costs.

Randomised control trials (RCTs) in the northern hemisphere (4, 6, 8, 12) and one in New Zealand(7) demonstrate that improving the thermal comfort of homes has generally positive effects of health and well-being, especially for certain populations (children, elderly) and certain health conditions (mental health, respiratory). There is a significant absence of Australian literature and related evidence on the relationship between housing conditions and health. This study addresses this gap.

The Victorian Healthy Homes Program (VHHP) delivered home thermal comfort and energy efficiency upgrades to 1000 vulnerable households in Victoria, Australia. An upgrade of up to AU$3,500 per home was fully paid for by the Victorian Government through Sustainability Victoria (SV). The program design incorporated a RCT to assess the impact of home upgrades on thermal comfort, well-being, health, energy use and costs to society.

The objectives of this paper are to report the (1) energy benefits; (2) health benefits; and (3) healthcare costs, of the VHHP. The trial protocol was published in 2022 (13).

## METHODS

Findings are reported in accordance with the CONSORT guidelines(14, 15).

### Trial design

This study was a staggered parallel group RCT design, where households were randomly assigned to one of two groups. Participants were recruited into the study over a three-year period from 2018 to 2020, inclusive. All households received the home upgrade either prior to winter (intervention group) or after winter (control group). The trial governance structure was that the program was funded and administered by Sustainability Victoria, it was delivered by the Australian Energy Foundation and the research component was independently conducted by the University of Technology Sydney.

### Change to trial design

As a result of the COVID-19 lockdown laws in Victoria, the program, and specifically the upgrades work in 2020, was significantly disrupted. Some intervention households did not receive their intervention prior to the winter of their recruitment year. For this reason, the following changes to the study were made (1) the 2020 post-winter visits were conducted via telephone, (2) the cost for remaining upgrades was reduced to a target average of AU$2,600 from December 2020.

### Participants and study setting

1000 households were recruited: 800 from western Melbourne (metropolitan) and 200 from the Goulburn Valle (regional). These areas were selected based on social or economic disadvantage and less favourable health outcomes. The average daily temperatures in these areas ranged from 3.4°C – 13.3°C (16). The eligibility criteria for participants are reported in the protocol paper (13).

Recruitment for the study occurred via the nine participating local government areas (LGAs) who disseminated promotional materials about the program to potential eligible householders within their jurisdiction. Interested householders completed an Expression of Interest and were then contacted by telephone by AEF staff to assess their eligibility and willingness to participate, and then to arrange a home visit with an interviewer. At this stage, vulnerability of the primary householder was assessed, which was defined as the participant being of low income i.e., having one of the Australian Government welfare cards, or receiving home care support services through local council or community organisations. During the first home visit and prior to commencing the interview, informed written consent was also sought from study participants to gain access to their energy and administrative health data. Consent forms were stored securely and separately from any other participant data to ensure confidentiality.

### Interventions

Based on a Victorian Residential Efficiency Scorecard (VRES)(17) assessment of the home quality and consultation with the participant, home upgrades were selected from a suite of interventions to improve thermal comfort within the budget, considering the Australian context of often poorly insulated houses and ease of installation with the least possible disruption to occupants. The range of home upgrades could include: ceiling and underfloor insulation, draught sealing external doors, space heating (new reverse cycle air conditioning or replacement of gas heater), changed downlights, and internal window coverings (see (13) for full list and description). Each home was reassessed under VRES after the upgrades, and the change in rating (on an overall scale of 10 stars) is the change in energy cost to run the home.

### Outcomes

#### Primary

The primary outcome was the average difference in measured indoor temperature between the intervention and control groups over winter (Table 1). Exposure to cold, measured by the time spent at indoor temperatures below 18^0^C (WHO recommended minimum), was also assessed (18). Winter was defined as the period from 22 June to 21 September, in line with the astronomical winter in Victoria.

**Table 1.** Outcome measures for the VHHP.

| **Data** | **Description** | **Data source** |
| --- | --- | --- |
| **Primary outcome** | Average daily temperature within the home | 30 min interval readings from data logger during winter |
|  | Exposure to cold conditions (number hours) | 30 min interval readings from data logger during winter |
| **Secondary (household level) outcome** | Change in average daily humidity within the home | 30 min interval readings from data logger during winter |
|  | Household energy costs | Self-reported survey |
|  | Total daily household energy consumption | Electricity and gas distributors |
| **Secondary (individual level) outcomes** | Health-related quality of life including health utilities | Self-reported surveys (SF-36, EQ-5D-5L and ASCOT) |
|  | Healthcare utilisation: |  |
|  | GP visits<br>Specialist visits<br>Diagnostic Tests | Medicare data (Services Australia) |
|  | Medicines prescribed | Pharmaceutical Benefits Scheme (PBS) data (Services Australia) |
|  | Hospital admissions and length of stay | Victorian Admitted Episodes Dataset (VAED) |
|  | Emergency Department presentations | Victorian Emergency Minimum Dataset (VEMD) |
|  | Respiratory Symptoms | Self-reported survey |
|  | Absenteeism | Self-reported survey |

Due to loss of battery power in some logger devices, 78 of the 250 households in the 2019 cohort reported varying levels of missing temperature and humidity data. The data missingness levels for these 78 households averaged around 30%. Data imputation was developed and utilised to address this issue(19).

#### Secondary

This study evaluated a number of secondary outcomes (Table 1). Secondary outcomes were assessed for all participating members in the household. The full set of secondary outcomes are reported in the protocol paper (13).

#### Quality of life

Quality of life data were collected using three established instruments, the EuroQol 5 dimension 5 level (EQ-5D-5L)(20-22), the short-form (SF)-36(23-25) and the Adult Social Care Outcomes Toolkit (ASCOT)(26). Participants completed all 3 questionnaires twice, before and after winter. The EQ-5D-5L and SF-36 were used to measure differences in health-related quality of life and the ASCOT for differences in social care outcomes(27).

#### Gas and electricity consumption

Gas and electricity consumption data obtained directly from the relevant companies were analysed for each home to determine changes in energy use and cost savings.

#### Health service utilisation and costs

Healthcare use and costs for individuals were identified from linked administrative health data extracts from the Medicare Benefits Schedule (MBS), Pharmaceutical Benefits Scheme (PBS), Victorian Admitted Episodes Dataset (VAED) and Victorian Emergency Minimum Dataset (VEMD). The pharmaceutical data includes all medicines listed on the PBS and dispensed to patients at a government-subsidised price. The MBS contains information on medical services that are subsidised by the Australian government including the benefits paid, the out-of-pocket costs to the patient as well as information about the nature of claim, and the service provider. The VAED contains International Classification of Diseases 10^th^ Revision ICD-10 and diagnosis related groups (DRG) codes on all admitted episodes from Victorian public and private acute hospitals. The VEMD contains demographic, administrative, and clinical data about presentations at Victorian public hospitals with designated emergency departments (EDs). A cost was assigned to each admitted hospital episode based on DRG and to each non-admitted ED visit based on urgency related group (URG) using related costs from the National Hospital Cost Data Collection (NHCDC) (ref). All costs were in Australian dollars (AUD) and adjusted to the 2020 year.

Data were extracted for the three years before and up to one year after the winter following recruitment. Data were analysed to establish the differences between the control and intervention groups in usage and cost.

#### Other secondary outcomes

The mMRC dyspnoea scale was used to record respiratory symptoms as it is a simple measure of breathlessness. Absenteeism was specified in the protocol as an important secondary outcome. In the after-winter surveys, we asked about days absent from (a) work, (b) study, and (c) usual activities for all adults in the household. Given the average age of our sample (76 years), there were very few participants engaged in work (99.5% indicated they had 0 days absent from work) or study (97.6% were not absent from study). The total number of days absent from usual activities over the winter period was summed and used as the measure of absenteeism.

### Sample size

A total of 984 households were included in the RCT which provides sufficient power to estimate effects for primary and secondary endpoints. All households were used in the analysis of both primary and secondary outcomes (unless consent was withdrawn during the study). Sample size calculations were based on two study endpoints, one household measure (indoor temperature) and one individual measure (the SF-36 MCS score). This was because the sample size needed for the primary outcome was significantly smaller than the secondary outcomes and we wanted to ensure we could detect significant differences in both outcomes. For the primary outcome, 125 households per group were required (power 90%, 15% loss, 5% significance level). For the secondary outcome, 475 households (950 participants) per group were required (80% power, 5% significance level, 20% loss, 2 individuals per household). For more details on these calculations see (13).

### Interim analysis and stopping guidelines

An interim blinded threshold analysis of the data occurred in October 2021 to provide feedback to Sustainability Victoria on the likely magnitude of effect needed to obtain outcomes based on current data. There were no group comparisons made at that point.

### Randomisation

The randomisation sequence was a 1:1 scheme stratified by LGA using random permuted blocks and randomly varying block size of 2 and 4. It was created at UTS using the Ralloc command in Stata 15.0 (28). The householders and the delivery partner (AEF) were informed about the study arm to which a household was assigned to after participant consent provision and collection of baseline data. (including home assessment). After data collection was complete, the random allocation was provided to analysts in a coded form so that primary and secondary analyses were conducted blinded to group allocation.

### Blinding

The trial was single blinded; we were unable to blind the households from the timing of the home upgrade. All ITT and PP analysis was conducted with group assignment in coded form only so that analysts were blinded to the household’s intervention status. Unblinding occurred after all analysis of the primary and secondary outcomes was complete.

### Statistical Methods

A model was developed that aggregated the outcome of interest over the entire winter period from 22 June to 21 September of the relevant year of recruitment, except for quality of life which was measured at 2 time-points only (before and after winter). The model specification is:

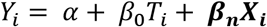

where Y is an outcome of interest for participant/household i, T denotes treatment group and *X*_*i*_signifies a number of covariates including household level variables for the primary and household level secondary outcomes, and including the individual’s cohort, age, gender and LGA for individual secondary outcomes. The outcomes of interest are described in Table 1. Where there is a before and after winter measure we control for the baseline values with variable “baseline” (Table 2). *β*_0_represents the difference between the control and intervention group. In the context of an RCT *β*_0_ can be interpreted as the impact of the VHHP. The standard errors were clustered at the household level for individual measures. All analyses included adjustment for LGA (stratifying variable for randomisation) and the year of recruitment (as the severity of the winter may affect the impact of the home upgrades).

**Table 2.**
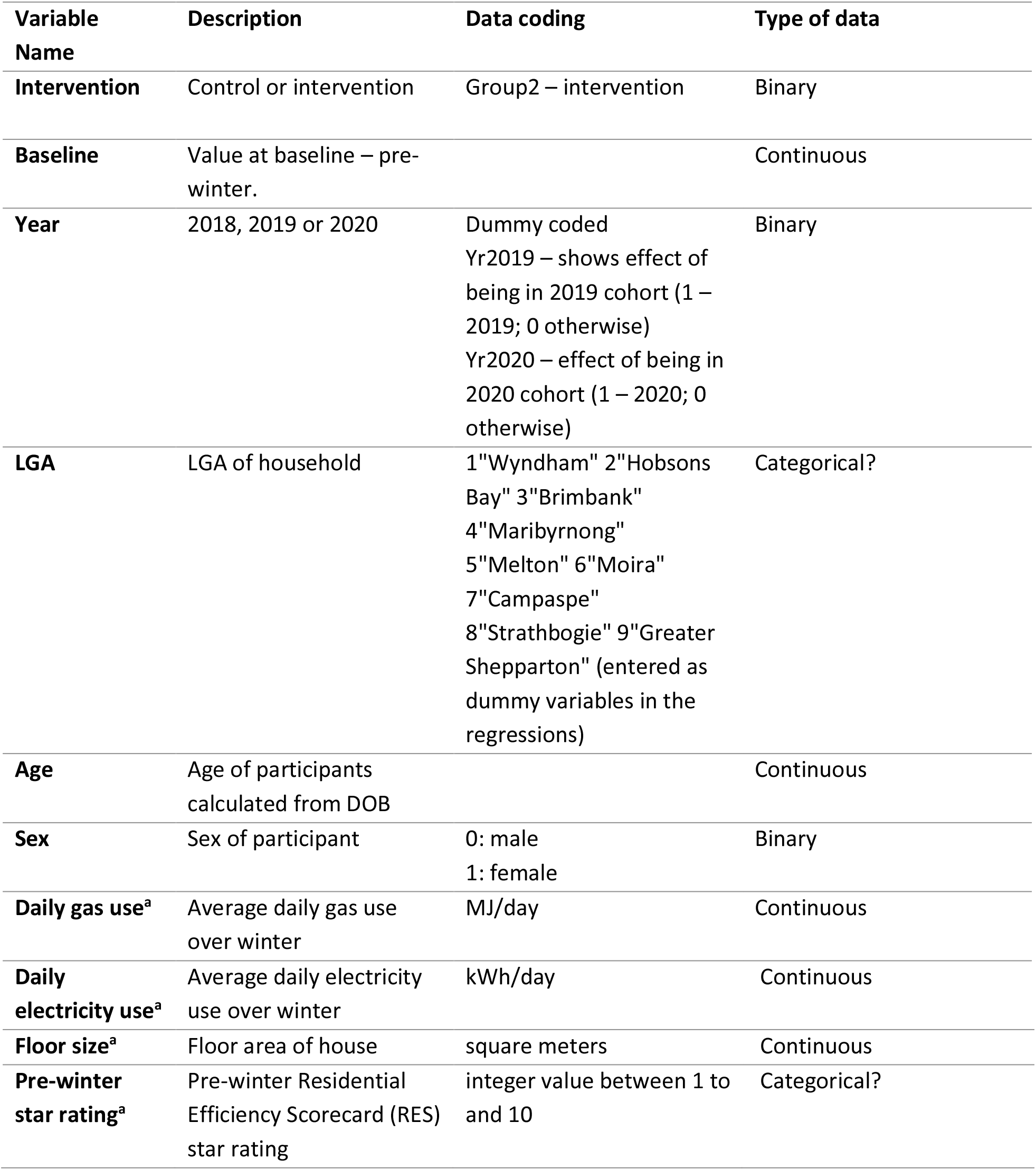
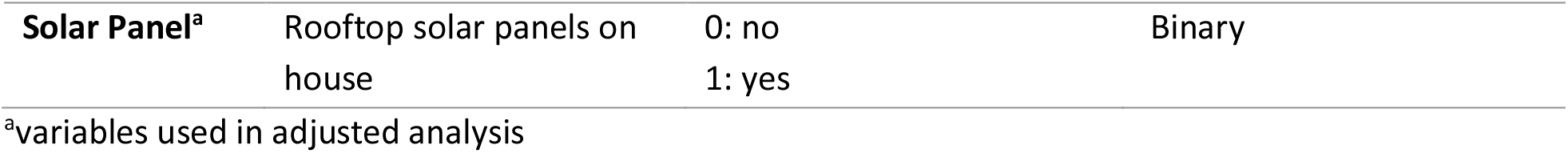
Control Variables, descriptions and coding in primary outcome adjusted and unadjusted regression models.

| **Variable Name** | **Description** | **Data coding** | **Type of data** |
| --- | --- | --- | --- |
| **Intervention** | Control or intervention | Group2 – intervention | Binary |
| **Baseline** | Value at baseline – pre-winter. |  | Continuous |
| **Year** | 2018, 2019 or 2020 | Dummy coded<br>Yr2019 – shows effect of being in 2019 cohort (1 – 2019; 0 otherwise)<br>Yr2020 – effect of being in 2020 cohort (1 – 2020; 0 otherwise) | Binary |
| **LGA** | LGA of household | 1"Wyndham" 2"Hobsons Bay" 3"Brimbank" 4"Maribyrnong" 5"Melton" 6"Moira" 7"Campaspe" 8"Strathbogie" 9"Greater Shepparton" (entered as dummy variables in the regressions) | Categorical? |
| **Age** | Age of participants calculated from DOB |  | Continuous |
| **Sex** | Sex of participant | 0: male<br>1: female | Binary |
| **Daily gas use<sup>a</sup>** | Average daily gas use over winter | MJ/day | Continuous |
| **Daily electricity use<sup>a</sup>** | Average daily electricity use over winter | kWh/day | Continuous |
| **Floor size<sup>a</sup>** | Floor area of house | square meters | Continuous |
| **Pre-winter star rating<sup>a</sup>** | Pre-winter Residential Efficiency Scorecard (RES) star rating | integer value between 1 to and 10 | Categorical? |
| **Solar Panel<sup>a</sup>** | Rooftop solar panels on house | 0: no<br>1: yes | Binary |

<sup>a</sup>variables used in adjusted analysis

The analysis described above may not fully capture the precise impact of intervention. Unlike a typical RCT conducted in the clinical setting where the impact lies on a linear scale (effect or no effect), participants in this study can choose to take the benefit of the intervention (home upgrade) as either improving thermal comfort (indoor temperature) or a reduction in their energy consumption (gas and electricity use). This can obscure the effects if only a unadjusted model is used. We therefore conduct a regression model that captures this unique the relationship between energy, indoor temperature, and other factors that mediate the relationship between the two key factors. Intervention dummy variable is further added into this model to determine its marginal effect, as shown in the Supplementary Material Table 3. This adjusted model is used for all outcomes linked to the primary outcome.

Two types of analysis were undertaken: (1) ITT analysis, which used all households allocated to the control and intervention groups, irrespective of whether they actually received the home upgrade intervention before winter (21 June); and (2) PP analysis, which includes only those households (individuals) who “completed” the treatment as originally allocated. Our definition of “completed” was that the intervention household must have received their full upgrade (all components) prior to the winter period in the year of recruitment, and control households must not have received any upgrade work prior to the end of winter (defined by 22 September) of the relevant year.

A range of modelling approaches were used for analysis. The logit model was used to analyse binary data; negative binomial models were used for count data and Ordinary Least Squares was used for continuous data.

Table 2 presents a list of control variables; the name refers to the name seen in the output of the regression analysis. Only the key variables are included here. Variables specific to each dataset are described in the relevant results sections below.

## RESULTS

### Numbers randomised

Figure 1**Error! Reference source not found**. shows the recruitment pathway for the VHHP. Of the 1,999 households contacted for eligibility, 984 were included in the study and randomised, 493 to the control group and 491 to the intervention group. These households are included in the ITT analysis.

### Losses and exclusions

Of the 984 households included in the trial (493 control; 491 intervention), 66 were lost to follow-up, 17 withdrew their consent and 14 had unusable data (data loggers not returned or lost). Due to the COVID-19 pandemic, 220 households did not receive their intervention during the protocol time frame. Seven hundred and sixty-four (764) households were included in the PP analysis (488 control 276 intervention). There were 1313 individuals (649 control: 664 intervention) for the full ITT analysis. For the PP analysis were 1015 individuals (641 control 374 intervention).

### Recruitment

The study commenced in February 2018. Recruitment commenced in January of each year, after the previous year’s after-winter interviews were completed. The program ended in May 2022 after the final control household upgrade was completed. The final administrative data was received in May 2023. Other final data (data loggers and surveys) arrived at various points throughout the trial between January 2021 and January 2022.

### Baseline data

Baseline characteristics for the control and intervention groups (ITT) are in Table 3. At both the household and individual level the control and intervention groups are not statistically different from each other on all the relevant variables and therefore the randomisation achieved the necessary balance.

**Table 3:**
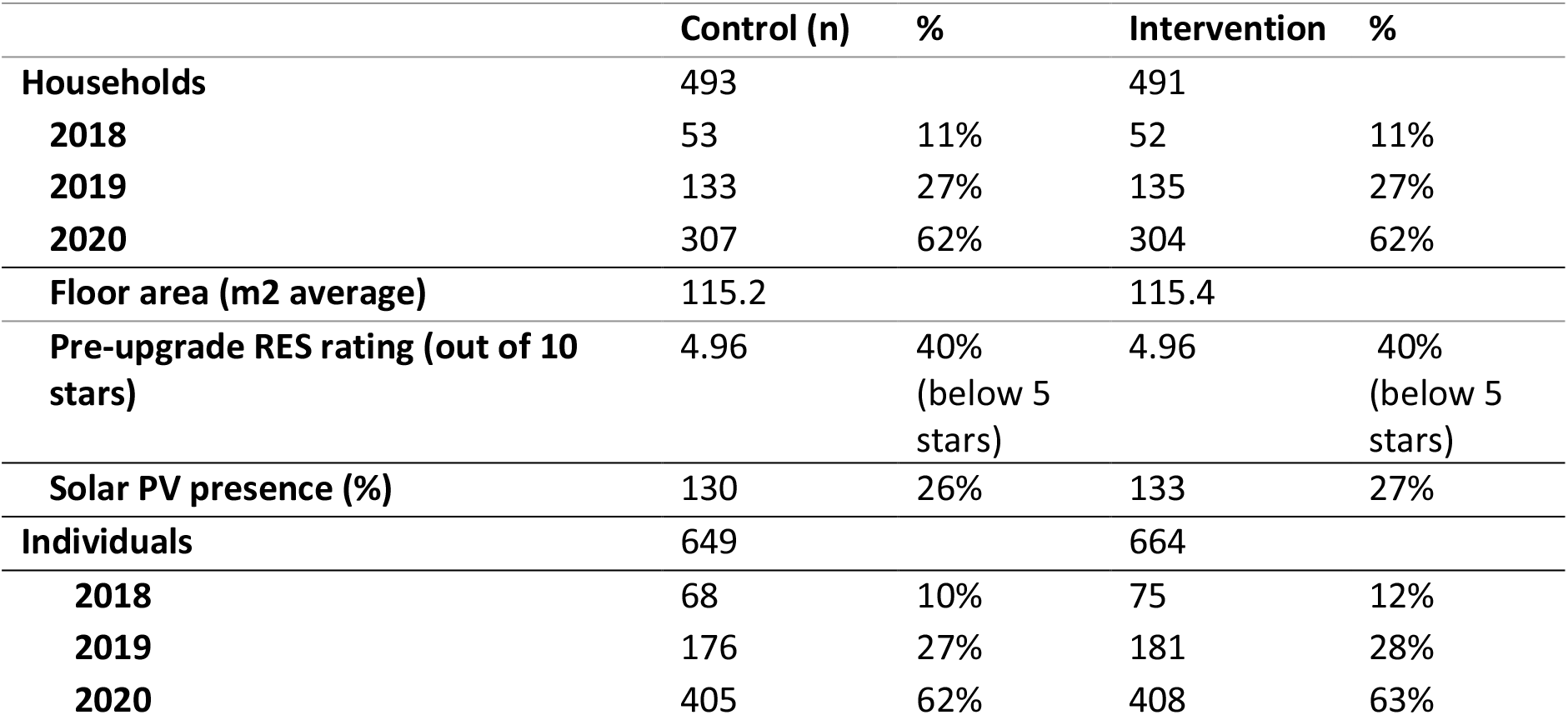
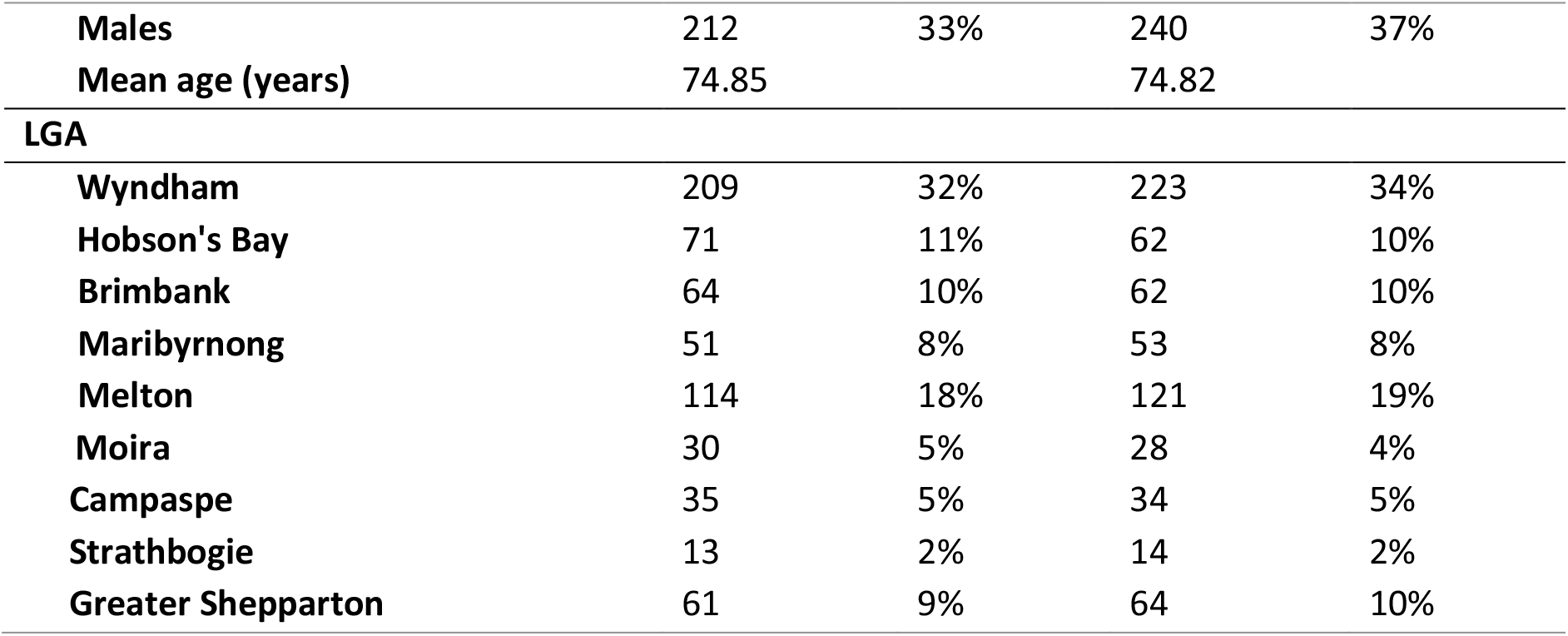
Baseline data for control and intervention groups.

### Outcomes and estimation

Table 4 presents the mean of all the outcomes for the control and intervention group and the raw differences, and the differences from the regression analysis (ITT). The full regression tables (ITT) are presented in supplementary material. The full regression tables for PP analysis are reported in the supplementary material only when significantly different from ITT results (Supplementary Material).

**Table 4:** Summary of energy and health regression results (ITT)

| **Outcome** | **Control Group**<br>**Mean** | **Intervention**<br>**Group**<br>**Mean** | **Raw mean difference**<br>**& 95%CI (t-test)** | **Difference from**<br>**regression model**<br>**(95% CI)** |
| --- | --- | --- | --- | --- |
| **Energy data** |  |  |  |  |
| **Unadjusted mean**<br>**winter**<br>**temperature (°C)** | 18.24 | 18.33 | 0.09<br>(-0.222, 0.390) | 0.09<br>(-0.217, 399) |
| **Mean winter**<br>**temperature<sup>a</sup> (°C)** | 18.24 | 18.33 | 0.09<br>(-0.222, 0.390) | 0.33<br>(0.05 0.60) |
| **Morning**<br>**temperature<sup>a</sup> (°C)** | 17.18 | 17.38 | 0.20<br>(-0.172, 0.585) | 0.47<br>(0.10 0.84) |
| **Hours/day below**<br>**18°C<sup>a</sup>, winter**<br>**period** | 11.77 | 11.43 | -0.34<br>(-1.16, 0.48) | -0.71<br>(-1.46 0.03) |
| **Hours/day below**<br>**hazardous**<br>**conditions<sup>a</sup>,**<br>**winter period** | 2.30 | 1.91 | -0.39<br>(-0.95, 0.17) | -0.925<br>(-1.60, -0.25) |
| **Perceived thermal**<br>**comfort (change**<br>**pre-post winter)** | -0.02 | 0.35 | 0.37<br>(0.16, 0.57) | 2.34 (1.83 3.01)<br>(OR) |
| **Gas use, MJ/day** | 222.00 | 202.00 | -20<br>(-40.47 0.93) | 25.49<br>(-43.05 -7.92) |
| **Gas use, entire winter, MJ** | 20424.00 | 18584.00 | -1840<br>(-3722.92 85.61) | -2345.08<br>(-3960.6 -728.64) |
| **Electricity use, KWh/day** | 13.78 | 14.32 | 0.54<br>(-1.32, 2.39) | 0.06<br>(-1.73, 2.88) |
| **Electricity use, entire winter (KWh)** | 1267.76 | 1317.44 | 49.68<br>(-121.44, 219.88) | -86.48<br>(-214.36 41.4) |
| **Administrative health data** |  |  |  |  |
| **MBS Services** | 14.11 | 12.7 | -1.041 (-.201,3.014) | 0.906<br>(0.813,1.011) (IRR) |
| **GP Services** | 2.58 | 2.62 | -0.045 (-0.367,0.277) | 1.016<br>(0.897,1.153) (IRR) |
| **PBS Services** | 16.17 | 16.53 | -0.0359 (-1.518,0.799) | 1.019<br>(0.946,1.097) (IRR) |
| **Hospital admissions** | 0.618 | 0.503 | -0.115 (-0.477 0.247) | 1.047 (0.750<br>1.463) (IRR) |
| **Hospital length of stay** | 1.081 | 1.103 | 0.022 (-0.494, 0.537) | 1.131 (0.716<br>1.786) (IRR) |
| **ED admissions** | 0.229 | 0.217 | 0.012 (-0.063, 0.087) | 1.030 (0.737<br>1.440) (IRR) |
| **After winter SF-36 MCS** | 43.981 | 46.005 | 2.025<br>(0.305, 3.744) | 1.730<br>(0.207, 3.254) |
| **After winter EQ-5D-5L utility score** | 0.605 | 0.624 | 0.019<br>(-0.022, 0.060) | 0.009<br>(-0.025 ,0.043) |
| **After winter ASCOT utility score** | 0.768 | 0.799 | 0.031<br>(0.010, 0.052) | 0.024<br>(0.006, 0.042) |
| **Survey Outcomes** |  |  |  |  |
| **Absenteeism** | 7.28 | 5.36 | -1.92 | 0.802 (0.536,1.20)<br>(IRR) |
| **Respiratory symptoms (MRCscore)** | -0.0566 | 0.188 | -0.245 (-0.388, -0.102) | -0.374 (-0.61, -<br>0.152) |

<sup>a</sup> refers to indoor temperature
Abbreviations: OR: Odds ratio. IRR: Incidence rate ratio. CI: Confidence interval.

### Primary outcome: Thermal comfort

#### Mean indoor temperature

Using the unadjusted analysis, home upgrades did not have an impact on mean indoor temperature over winter (0.091 °C; 95% CI -0.217,0.399) for both ITT and PP analyses (Supplementary Material Table 1 and 2). Under the adjusted model, home upgrades did have statistically significant impact on mean indoor temperature over winter (0.326 °C; 95% CI 0.047, 0.605) for the ITT analysis.

The intervention was also assessed by time of day, mornings (8am-12pm), afternoons (12pm-5pm), evenings (5pm-10pm), and overnight (10pm-8am) (Supplementary Material Tables 4 to 7). Home upgrades had the largest impact on indoor temperature in the mornings, increasing indoor temperature by 0.47°C (95% CI 0.105, 0.836) for the ITT analysis.

#### Exposure to cold

Home upgrades had a positive and statistically significant impact, reducing exposure to cold indoor conditions analysis (−0.71 hours/day (43 mins); 95% CI -1.456 0.029) for the ITT analysis. This effect was larger for the PP analysis (−0.93 hours/day (56 mins); 95% CI -1.813 -0.056) (Supplementary Material Table 8 and 9).

Assessment of differences in exposure to cold indoor conditions was not significant for mornings in ITT analysis (−0.16 hours/day (10 mins); 95% CI -1.898, 0.058), however the effect was significant for PP analysis (−0.22 hours/day (13 mins); 95% CI -0.416, -0.031). This effect is equivalent to ∼9.6% reduction in exposure to cold during the morning, given that the entire group experiences on average 2.3 hours (138 mins) out of the 4 morning hours below 18°C. Home upgrades did not impact afternoon hours below 18°C in ITT analysis (−0.14 hours/day (9 mins); 95% CI -0.346, 0.059), but was significant in PP analysis (−0.26 hours/day (16 mins); 95% CI -0.504, -0.012) (Supplementary Material Tables 10 to 17).

### Secondary outcomes

#### Perceived thermal comfort

Home upgrades were found to improve household’s perceived thermal comfort, in line with findings on measured thermal comfort, where intervention households were 2.34 times (95% CI 1.83 3.01) more likely to report an increase in self-reported thermal comfort compared to control households for the ITT analysis. (Supplementary Material Table 18)

#### Humidity and hazardous conditions

Internal relative humidity levels were typically around 50%. High humidity levels, in conjunction with low indoor temperature, can lead to respiratory hazards. The WHO benchmarks a combination of relative humidity over 65% and temperatures below 16°C as hazardous to health (29). ITT results showed that home upgrades led to nearly an hour (55 minutes) of reduced exposure to hazardous conditions per day (−0.925 hours/day, 95% CI -1.599 -0.250) (Supplementary Material Table 19).

#### Energy Outcomes

##### Gas and electricity use

Home upgrades did not have an impact on average electricity use (−0.943 kWh/day, 95% CI 0.445, - 2.231). This is not surprising because gas heating dominates in Victoria, with 74% of study households using gas as main heating source. Upgrades did have a significant impact on gas use with less gas use in the upgraded homes (−7.08 kWh/day, 95% CI -11.959 -2.201) (Supplementary Material Tables 20 and 21).

##### Energy costs

We used the average Victorian Default Offer electricity rate (AU$0.29/kWh) and gas rate (AU$0.107/kWh) to quantify the financial impact of gas usage savings in intervention households, which was 7.08 kWh (25.49 MJ) per day and equates to $AU69.70 in savings per winter per household.

#### Quality of life

Two quality of life measures show significant differences between the control and intervention group (Supplementary Material). The SF-36 MCS score after winter is significantly different for the intervention group compared to the control group (1.73, 95% CI: 0.21, 3.25) (Supplementary Material Table 22) such that they showed a significantly greater improvement in mental health. The SF36 PCS score was not significant (Table 23 Supplementary Material). The ASCOT scores after winter are also significantly higher for the intervention group in the ITT analysis (0.024, 95% CI: 0.006, 0.042) (Table 24 Supplementary Material). There were no significant differences between the control and intervention groups for EQ-5D-5L or ‘health today’ scores after winter (Supplementary Material Tables 25 and 26).

#### Healthcare utilisation and cost

Overall group differences in both MBS, PBS and GP service use were not statistically significant but there was a trend that the intervention group used fewer medical services over winter (12.8 services: intervention group; 15.1 services control group).

When looking at costs the differences between control and intervention group in total cost for private medical and diagnostic services (called MBS charge) was significant (AU$-156.57; 95% CI: - 310.68, -2.47) such that the intervention group incurred significantly fewer total service costs over winter. In 2020 there was lower service use and hence overall costs compared to the 2018 and 2109, which is likely attributable to reduced use during COVID-19 related lockdowns. All regression results are shown in Tables 27-30 Supplementary Material.

A negative binomial regression model showed no significant difference between the control and intervention groups for pharmaceutical services, gross price, net benefits or patient contributions. All regression results are shown in (Tables 31-34 Supplementary Material).

There was no significant difference between the control and intervention groups for the outcome of the number of hospitalisations, hospital costs, length of stay (days in hospital) and emergency department visits. An overall trend of lower hospitalisations and hospital costs was observed for the intervention group. All regression tables are shown in Table 35-39 (Supplementary Material).

Total healthcare costs were defined as the sum of all healthcare service use costs during the 3-month winter period. These data are based on costs as reported in the MBS, PBS and hospital datasets. the regression analysis predicting total healthcare costs and shows that the intervention group on average uses AU$886.51 (CI: -1879.25-106.23) less of healthcare costs than the control group (Table 41 Supplementary Material).

#### Other secondary outcomes

The control group had a higher number of days absent from usual activities (mean = 7.3, SD 15.8) than the intervention group (mean = 5.4, SD 13.3). This group difference was not significant in ITT analysis (coefficient = -0.22; 95% CI -0.62, 0.18; p=0.28). It was stronger in PP analysis (coefficient = - 0.46; 95% CI -0.94, 0.02; p=0.058) (Tables 42-43 Supplementary Material).

Respiratory symptoms were measured by the self-reported changes in mMRC dyspnoea scale. Logistic regression indicated that individuals in the intervention group had a reduction (improvement) in mMRC score relative to those in the control group over winter (coefficient = -0.38; 95% CI -0.61, -0.15; p=0.001).

## DISCUSSION

The VHHP is the first comprehensive evaluation of the thermal comfort, energy use and health impacts of home upgrades to be conducted alongside a randomised controlled trial in Australia. The overall results indicate that home upgrades provide important benefits in thermal comfort and quality of life, and in reduction in energy costs.

The increase in mean indoor temperatures by an average of 0.33°C and reduced exposure to unhealthy indoor conditions by an average of 43 mins/day correlate to valuable energy savings and improved comfort for participants.

These results concur with international evidence particularly in relation to energy benefits but also for mental health improvements. Our study demonstrates that health and energy benefits depend on baseline housing conditions with poorer quality houses benefiting more. This suggests careful targeting of the intervention is important to maximize benefits, similar to (30, 31).

A major factor for both the conduct and outcomes of this study was the impact of the COVID-19 pandemic. First, there was an impact on the per-protocol sample size for the 2020 cohort and consequently a loss of power to detect significant differences. There was a very large percentage of households in the intervention group that did not receive their upgrade in time. Despite this we show significant effects in some outcomes. Consequently, we believe our effect estimates are likely a large underestimate of the true impact of home upgrades. This is even more so when we consider that we are constraining our analysis to 3 months over winter. The effects of these upgrades endure for years and these benefits are not captured in the current analysis.

Second, the 2020 cohort effect is significant in many of the regression models. The most obvious example of the “2020” pandemic cohort effect is evident in the administrative health data. We observe that there is a reduction in healthcare service use and cost and our survey data reveals that individuals spent more time at home. They also are likely to use their energy differently because of these changes in behaviour. Our post winter 2020 survey was modified to capture some of these effects.

In this study, we did not control for the exact timing of the intervention and therefore assume that the “length of the intervention dose” is the same for all intervention participants (3 months over winter). However, we acknowledge that households received their upgrades at differing points in time in the pre-winter period and over differing durations due to the practicalities of scheduling the various trades to undertake the works. This means that some households have had the “dose” of the intervention longer than others.

This evaluation has yielded valuable insights into the challenges associated with undertaking population interventions of this nature. In particular, because the intervention requires modifications to the participants’ homes, there are important occupational health and safety issues that require careful planning and consideration. The design of the trial also required that the modifications be conducted with time constraints, and the ability to control this proved to be challenging and beyond control of the program.

Despite these challenges, the program provides rigorous, novel evidence that modest home upgrades can have important impacts on thermal comfort, energy use healthcare costs and wellbeing. The strengths of this study include the randomised deign, comprehensive objective and subjective thermal comfort data, objective linked health data and quality of life information for the same cohort. These feelings add much needed evidence to the research base on the co-benefits of home upgrades.

## OTHER INFORMATION

### REGISTRATION

Australian and New Zealand Clinical Trials Registry: ACTRN12618000160235; Pre-results

### PROTOCOL

The protocol for this trial was published in BMJ Open in 2022. The full reference is:

Campbell M, Page K, Longden T, Kenny P, Hossain L, Wilmot K, Kelly S, Kim Y, Haywood P, Mulhern B, Goodall S. Evaluation of the Victorian Healthy Homes Program: protocol for a randomised controlled trial. BMJ open. 2022 Apr 1;12(4):e053828.

## Supporting information

Supplementary Files VHHP

## Data Availability

Data are held in a Secure research environment (SURE) and are archived. They are not available to other researchers because of the consent and ethics required by the various Government authorities who manage health data.

## ETHICS AND DISSEMINATION

Ethical approval was received from Victorian Department of Human Services (DHS) Human Research Ethics Committee, University of Technology (UTS) Sydney Human Research Ethics Committee and Australian Government Department of Veterans’ Affairs (DVA).

The Victorian Healthy Homes Program is funded by the Victorian State Government through the Sustainability Victoria Fund through Sustainability Victoria

## AUTHOR’S CONTRIBUTIONS

KP, LH, DL, KW, YK were responsible for drafting and critically revising the manuscript. KVG, RV, TC and MC also critically revised the manuscript. Data extraction and quality assessment was performed by PK, KP, DL, LH, KW, YK. Authors KP, PK, DL, LH, YK, KW, KVG and RV are responsible for the data analysis. All authors contributed to the implementation of the study protocol. All authors have critically reviewed the manuscript and approved the final submission.

VHHP team members (alphabetically): Jay Falletta^2^, Stephen Goodall^1^, Phil Haywood^1^, Deepu Krishnan^2^, Brendan Mulhern^1^, Matthew Soeberg^3^.

^1^Centre for Health Economics Research and Evaluation, University of Technology Sydney, Australia ^2^Institute for Sustainable Futures, University of Technology Sydney, Australia, ^3^Sustainability Victoria, Melbourne, Australia.

## ACKNOWLEDGEMENTS

The Victorian Healthy Homes Program is household energy efficiency program targeted at lower-income households and is funded by the Victorian Government. We thank the Commonwealth DHS (Services Australia) for enabling access to consented records from the Medicare Benefits Schedule and the Pharmaceutical Benefits Schedule. We also thank the Victorian Department of Health and Human Services for providing access to consented records for admitted and non-admitted hospital episodes of care, emergency department presentation episodes, and mortality data. We thank Victorian Electricity and gas distributors for providing access to household energy consumption data. We also thank the Centre for Victorian Data Linkage for conducting record linkage. We also acknowledge and thank all households and individuals who participated in the Victorian Healthy Homes Program and who consented for their data to be used for research and evaluation purposes. We thank the many staff at SV and AEF for their commitment and diligence to the implementation of the VHHP without whom the work would not have been possible.

