## Supplementary Files VHHP for "Outcomes from the Victorian Healthy Homes Program: a randomised control trial of home energy upgrades"

Supplementary Material

Regression Tables

Table 1. Linear regression for Indoor temperature (ITT)

| **Number of obs** | = | **888** | | | | |
|---|---|---|---|---|---|---|
| **Marginal R-squared** | = | 0.003 | | | | |
| | Coef. | Std. Err. | t | P>\|t\| | [95% Conf. | Interval] |
| **Intervention** | 0.091 | 0.157 | 0.578 | 0.563 | -0.217 | 0.399 |
| **2019 Group** | -0.242 | 0.279 | -0.866 | 0.387 | -0.790 | 0.306 |
| **2020 Group** | 0.037 | 0.276 | 0.135 | 0.893 | -0.505 | 0.580 |
| **Intercept** | 18.284 | 0.267 | 68.430 | <0.001 | 17.759 | 18.808 |

*adjusted for LGA

Table 2 Linear regression for Indoor temperature (PP)

| **Number of obs** | = | **692** | | | | |
|---|---|---|---|---|---|---|
| **Marginal R-squared** | = | 0.007 | | | | |
| | Coef. | Std. Err. | t | P>\|t\| | [95% Conf. | Interval] |
| **Intervention** | 0.021 | 0.177 | 0.121 | 0.904 | -0.325 | 0.368 |
| **2019 Group** | 0.017 | 0.298 | 0.057 | 0.954 | -0.569 | 0.603 |
| **2020 Group** | 0.389 | 0.282 | 1.380 | 0.168 | -0.165 | 0.944 |
| **Intercept** | 18.056 | 0.266 | 67.903 | <0.001 | 17.533 | 18.578 |

*adjusted for LGA

Table 3 Adjusted linear regression for Indoor temperature (ITT)

| **Number of obs** | = | **662** | | | | |
|---|---|---|---|---|---|---|
| **Marginal R-squared** | = | 0.375 | | | | |
| | Coef. | Std. Err. | t | P>\|t\| | [95% Conf. | Interval] |
| **Intervention** | 0.326 | 0.142 | 2.298 | 0.022 | 0.047 | 0.605 |
| **Daily Gas Use (kWh)** | 0.031 | 0.002 | 16.583 | <0.001 | 0.027 | 0.035 |
| **Daily Elec Use (kWh)** | 0.080 | 0.007 | 11.049 | <0.001 | 0.066 | 0.094 |
| **Floor size (log sqm)** | -3.355 | 0.575 | -5.835 | <0.001 | -4.484 | -2.226 |
| **2019 Group** | 0.145 | 0.245 | 0.591 | 0.555 | -0.336 | 0.625 |
| **2020 Group** | 0.076 | 0.226 | 0.337 | 0.737 | -0.368 | 0.52 |
| **Pre-winter star rating** | 0.300 | 0.044 | 6.819 | <0.001 | 0.214 | 0.386 |
| **Solar Panel (Y/N)** | -0.621 | 0.186 | -3.343 | 0.001 | -0.986 | -0.256 |
| **Intercept** | 20.750 | 1.256 | 16.527 | <0.001 | 18.285 | 23.216 |

*adjusted for LGA

Table 4 Linear regression for Indoor temperature (ITT), morning

| **Number of obs** | = | **662** | | | | |
|---|---|---|---|---|---|---|
| **Marginal R-squared** | = | 0.313 | | | | |
| | Coef. | Std. Err. | t | P>\|t\| | [95% Conf. | Interval] |
| **Intervention** | 0.471 | 0.186 | 2.526 | 0.012 | 0.105 | 0.836 |
| **Daily Gas Use (kWh)** | 0.035 | 0.002 | 14.130 | <0.001 | 0.030 | 0.039 |
| **Daily Elec Use (kWh)** | 0.092 | 0.009 | 9.647 | <0.001 | 0.073 | 0.110 |
| **Floor size (log sqm)** | -4.506 | 0.755 | -5.970 | <0.001 | -5.988 | -3.024 |
| **2019 Group** | 0.499 | 0.321 | 1.555 | 0.120 | -0.131 | 1.130 |
| **2020 Group** | 0.993 | 0.297 | 3.344 | 0.001 | 0.410 | 1.575 |
| **Pre-winter star rating** | 0.190 | 0.058 | 3.294 | 0.001 | 0.077 | 0.304 |
| **Solar Panel (Y/N)** | -0.322 | 0.244 | -1.322 | 0.187 | -0.801 | 0.157 |
| **Intercept** | 21.472 | 1.648 | 13.025 | <0.001 | 18.235 | 24.709 |

*adjusted for LGA

Table 5 Linear regression for Indoor temperature (ITT), afternoon

| **Number of obs** | = | **662** | | | | |
|---|---|---|---|---|---|---|
| **Marginal R-squared** | = | 0.311 | | | | |
| | Coef. | Std. Err. | t | P>\|t\| | [95% Conf. | Interval] |
| **Intervention** | 0.271 | 0.159 | 1.696 | 0.090 | -0.043 | 0.584 |
| **Daily Gas Use (kWh)** | 0.030 | 0.002 | 14.130 | <0.001 | 0.026 | 0.034 |
| **Daily Elec Use (kWh)** | 0.075 | 0.008 | 9.289 | <0.001 | 0.060 | 0.091 |
| **Floor size (log sqm)** | -3.465 | 0.646 | -5.363 | <0.001 | -4.734 | -2.196 |
| **2019 Group** | 0.111 | 0.275 | 0.405 | 0.686 | -0.429 | 0.651 |
| **2020 Group** | 0.576 | 0.254 | 2.268 | 0.024 | 0.077 | 1.075 |
| **Pre-winter star rating** | 0.284 | 0.049 | 5.748 | <0.001 | 0.187 | 0.381 |
| **Solar Panel (Y/N)** | -0.560 | 0.209 | -2.682 | 0.007 | -0.970 | -0.150 |
| **Intercept** | 21.778 | 1.411 | 15.433 | <0.001 | 19.007 | 24.549 |

*adjusted for LGA

Table 6 Linear regression for Indoor temperature (ITT), evening

| **Number of obs** | **=** | **662** | | | | |
|---|---|---|---|---|---|---|
| **Marginal R-squared** | = | 0.275 | | | | |
| | Coef. | Std. Err. | t | P>\|t\| | [95% Conf. | Interval] |
| **Intervention** | 0.317 | 0.166 | 1.913 | 0.056 | -0.008 | 0.643 |
| **Daily Gas Use (kWh)** | 0.029 | 0.002 | 13.228 | <0.001 | 0.025 | 0.033 |
| **Daily Elec Use (kWh)** | 0.064 | 0.008 | 7.564 | <0.001 | 0.047 | 0.081 |
| **Floor size (log sqm)** | -2.512 | 0.675 | -3.722 | <0.001 | -3.838 | -1.187 |
| **2019 Group** | -0.131 | 0.287 | -0.458 | 0.647 | -0.694 | 0.431 |
| **2020 Group** | 0.173 | 0.324 | 0.534 | 0.593 | -0.463 | 0.809 |

| | | | | | | |
|---|---|---|---|---|---|---|
| **Pre-winter star rating** | 0.381 | 0.052 | 7.344 | <0.001 | 0.279 | 0.483 |
| **Solar Panel (Y/N)** | -0.906 | 0.218 | -4.157 | <0.001 | -1.334 | -0.478 |
| **Intercept** | 21.32 | 1.493 | 14.28 | <0.001 | 18.388 | 24.251 |

*adjusted for LGA

Table 7 Linear regression for Indoor temperature (ITT), overnight

| | | | | | | |
|---|---|---|---|---|---|---|
| **Number of obs** | = | **662** | | | | |
| **Marginal R-squared** | = | 0.369 | | | | |
| | Coef. | Std. Err. | t | P>\|t\| | [95% Conf. | Interval] |
| **Intervention** | 0.297 | 0.150 | 1.980 | 0.048 | 0.002 | 0.591 |
| **Daily Gas Use (kWh)** | 0.031 | 0.002 | 15.802 | <0.001 | 0.027 | 0.035 |
| **Daily Elec Use (kWh)** | 0.085 | 0.008 | 11.145 | <0.001 | 0.070 | 0.100 |
| **Floor size (log sqm)** | -3.290 | 0.607 | -5.419 | <0.001 | -4.483 | -2.098 |
| **2019 Group** | 0.165 | 0.258 | 0.638 | 0.523 | -0.342 | 0.672 |
| **2020 Group** | -0.533 | 0.239 | -2.234 | 0.026 | -1.002 | -0.065 |
| **Pre-winter star rating** | 0.306 | 0.046 | 6.579 | <0.001 | 0.214 | 0.397 |
| **Solar Panel (Y/N)** | -0.621 | 0.196 | -3.167 | 0.002 | -1.007 | -0.236 |
| **Intercept** | 19.719 | 1.326 | 14.870 | <0.001 | 17.116 | 22.323 |

*adjusted for LGA

Table 8 Linear regression for hours of Exposure to cold (ITT)

| | | | | | | |
|---|---|---|---|---|---|---|
| **Number of obs** | = | **662** | | | | |
| **Marginal R-squared** | = | 0.390 | | | | |
| | Coef. | Std. Err. | t | P>\|t\| | [95% Conf. | Interval] |
| **Intervention** | -0.713 | 0.378 | -1.886 | 0.060 | -1.456 | 0.029 |
| **Daily Gas Use (kWh)** | -0.082 | 0.005 | -16.471 | <0.001 | -0.092 | -0.072 |
| **Daily Elec Use (kWh)** | -0.224 | 0.019 | -11.642 | <0.001 | -0.262 | -0.187 |
| **Floor size (log sqm)** | 10.258 | 1.537 | 6.676 | <0.001 | 7.241 | 13.275 |
| **2019 Group** | 0.190 | 0.653 | 0.291 | 0.771 | -1.092 | 1.472 |
| **2020 Group** | -0.931 | 0.675 | -1.379 | 0.168 | -2.257 | 0.394 |
| **Pre-winter star rating** | -0.731 | 0.118 | -6.204 | <0.001 | -0.963 | -0.500 |
| **Solar Panel (Y/N)** | 1.051 | 0.496 | 2.118 | 0.035 | 0.077 | 2.026 |
| **Intercept** | 2.534 | 3.376 | 0.751 | 0.453 | -4.095 | 9.164 |

*adjusted for LGA

Table 9 Linear regression for hours of Exposure to cold (PP)

| | | | | | | |
|---|---|---|---|---|---|---|
| **Number of obs** | = | **512** | | | | |
| **Marginal R-squared** | = | 0.393 | | | | |
| | Coef. | Std. Err. | t | P>\|t\| | [95% Conf. | Interval] |

| | | | | | | |
|---|---|---|---|---|---|---|
| **Intervention** | -0.934 | 0.447 | -2.09 | 0.037 | -1.813 | -0.056 |
| **Daily Gas Use (kWh)** | -0.076 | 0.006 | -13.697 | <0.001 | -0.087 | -0.065 |
| **Daily Elec Use (kWh)** | -0.232 | 0.02 | -11.427 | <0.001 | -0.272 | -0.192 |
| **Floor size (log sqm)** | 10.221 | 1.683 | 6.074 | <0.001 | 6.915 | 13.527 |
| **2019 Group** | 0.059 | 0.723 | 0.082 | 0.935 | -1.361 | 1.479 |
| **2020 Group** | -1.576 | 0.69 | -2.284 | 0.023 | -2.932 | -0.220 |
| **Pre-winter star rating** | -0.644 | 0.13 | -4.952 | <0.001 | -0.900 | -0.389 |
| **Solar Panel (Y/N)** | 0.98 | 0.548 | 1.789 | 0.074 | -0.096 | 2.057 |
| **Intercept** | 2.321 | 3.67 | 0.632 | 0.527 | -4.889 | 9.532 |

*adjusted for LGA

Table 10 Linear regression for hours of Exposure to cold (ITT), mornings

| **Number of obs** | = | **662** | | | | |
|---|---|---|---|---|---|---|
| **Marginal R-squared** | = | 0.390 | | | | |
| | Coef. | Std. Err. | t | P>\|t\| | [95% Conf. | Interval] |
| **Intervention** | -0.159 | 0.084 | -0.324 | 0.005 | -1.898 | 0.058 |
| **Daily Gas Use (kWh)** | -0.015 | 0.001 | -0.017 | -0.013 | -13.353 | <0.001 |
| **Daily Elec Use (kWh)** | -0.042 | 0.004 | -0.050 | -0.033 | -9.753 | <0.001 |
| **Floor size (log sqm)** | 2.313 | 0.340 | 1.645 | 2.980 | 6.804 | <0.001 |
| **2019 Group** | -0.227 | 0.145 | -0.511 | 0.057 | -1.571 | 0.117 |
| **2020 Group** | -0.619 | 0.134 | -0.882 | -0.357 | -4.634 | <0.001 |
| **Pre-winter star rating** | -0.070 | 0.026 | -0.121 | -0.019 | -2.700 | 0.007 |
| **Solar Panel (Y/N)** | 0.099 | 0.110 | -0.117 | 0.315 | 0.901 | 0.368 |
| **Intercept** | -0.178 | 0.742 | -1.636 | 1.280 | -0.240 | 0.810 |

*adjusted for LGA

Table 11 Linear regression for hours of Exposure to cold (PP), mornings

| **Number of obs** | = | **512** | | | | |
|---|---|---|---|---|---|---|
| **Marginal R-squared** | = | 0.336 | | | | |
| | Coef. | Std. Err. | t | P>\|t\| | [95% Conf. | Interval] |
| **Intervention** | -0.223 | 0.098 | -2.276 | 0.023 | -0.416 | -0.031 |
| **Daily Gas Use (kWh)** | -0.014 | 0.001 | -11.229 | <0.001 | -0.016 | -0.011 |
| **Daily Elec Use (kWh)** | -0.043 | 0.004 | -9.598 | <0.001 | -0.051 | -0.034 |
| **Floor size (log sqm)** | 2.278 | 0.369 | 6.174 | <0.001 | 1.553 | 3.003 |
| **2019 Group** | -0.232 | 0.158 | -1.467 | 0.143 | -0.544 | 0.079 |
| **2020 Group** | -0.753 | 0.151 | -4.978 | <0.001 | -1.050 | -0.456 |
| **Pre-winter star rating** | -0.054 | 0.029 | -1.905 | 0.057 | -0.110 | 0.002 |
| **Solar Panel (Y/N)** | 0.082 | 0.120 | 0.684 | 0.494 | -0.154 | 0.318 |
| **Intercept** | -0.160 | 0.805 | -0.199 | 0.842 | -1.741 | 1.421 |

*adjusted for LGA

Table 12 Linear regression for hours of Exposure to cold (ITT), afternoon

| **Number of obs** | = | **662** | | | | |
|---|---|---|---|---|---|---|
| **Marginal R-squared** | = | 0.390 | | | | |
| | Coef. | Std. Err. | t | P>\|t\| | [95% Conf. | Interval] |
| **Intervention** | -0.143 | 0.103 | -1.389 | 0.165 | -0.346 | 0.059 |
| **Daily Gas Use (kWh)** | -0.019 | 0.001 | -14.277 | <0.001 | -0.022 | -0.017 |
| **Daily Elec Use (kWh)** | -0.044 | 0.005 | -8.393 | <0.001 | -0.055 | -0.034 |
| **Floor size (log sqm)** | 2.355 | 0.421 | 5.594 | <0.001 | 1.528 | 3.181 |
| **2019 Group** | 0.147 | 0.178 | 0.823 | 0.411 | -0.204 | 0.497 |
| **2020 Group** | -0.449 | 0.216 | -2.080 | 0.038 | -0.872 | -0.025 |
| **Pre-winter star rating** | -0.156 | 0.032 | -4.820 | <0.001 | -0.220 | -0.093 |
| **Solar Panel (Y/N)** | 0.202 | 0.136 | 1.486 | 0.138 | -0.065 | 0.469 |
| **Intercept** | -0.252 | 0.936 | -0.269 | 0.788 | -2.091 | 1.587 |

*adjusted for LGA

Table 13 Linear regression for hours of Exposure to cold (PP), afternoon

| **Number of obs** | = | **512** | | | | |
|---|---|---|---|---|---|---|
| **Marginal R-squared** | = | 0.307 | | | | |
| | Coef. | Std. Err. | t | P>\|t\| | [95% Conf. | Interval] |
| **Intervention** | -0.258 | 0.125 | -2.059 | 0.040 | -0.504 | -0.012 |
| **Daily Gas Use (kWh)** | -0.017 | 0.002 | -11.196 | <0.001 | -0.020 | -0.014 |
| **Daily Elec Use (kWh)** | -0.046 | 0.006 | -8.073 | <0.001 | -0.057 | -0.035 |
| **Floor size (log sqm)** | 2.159 | 0.471 | 4.584 | <0.001 | 1.233 | 3.084 |
| **2019 Group** | 0.152 | 0.202 | 0.752 | 0.452 | -0.245 | 0.550 |
| **2020 Group** | -0.580 | 0.193 | -3.003 | 0.003 | -0.960 | -0.201 |
| **Pre-winter star rating** | -0.148 | 0.036 | -4.076 | <0.001 | -0.220 | -0.077 |
| **Solar Panel (Y/N)** | 0.255 | 0.153 | 1.661 | 0.097 | -0.047 | 0.556 |
| **Intercept** | 0.096 | 1.027 | 0.094 | 0.925 | -1.922 | 2.114 |

*adjusted for LGA

Table 14 Linear regression for hours of Exposure to cold (ITT), evening

| **Number of obs** | = | **662** | | | | |
|---|---|---|---|---|---|---|
| **Marginal R-squared** | = | 0.390 | | | | |
| | Coef. | Std. Err. | t | P>\|t\| | [95% Conf. | Interval] |
| **Intervention** | -0.133 | 0.090 | -1.472 | 0.142 | -0.310 | 0.044 |
| **Daily Gas Use (kWh)** | -0.015 | 0.001 | -12.901 | <0.001 | -0.018 | -0.031 |
| **Daily Elec Use (kWh)** | -0.029 | 0.005 | -6.282 | <0.001 | -0.038 | -0.020 |
| **Floor size (log sqm)** | 1.253 | 0.368 | 3.401 | 0.001 | 0.530 | 1.977 |
| **2019 Group** | 0.254 | 0.156 | 1.629 | 0.104 | -0.052 | 0.560 |
| **2020 Group** | -0.155 | 0.209 | -0.744 | 0.457 | -0.565 | 0.254 |

| | | | | | | |
|---|---|---|---|---|---|---|
| **Pre-winter star rating** | -0.167 | 0.028 | -5.864 | <0.001 | -0.222 | -0.111 |
| **Solar Panel (Y/N)** | 0.199 | 0.119 | 1.676 | 0.094 | -0.034 | 0.432 |
| **Intercept** | 0.649 | 0.830 | 0.782 | 0.434 | -0.980 | 2.278 |

*adjusted for LGA

Table 15 Linear regression for hours of Exposure to cold (PP), evening

| **Number of obs** | = | **512** | | | | |
|---|---|---|---|---|---|---|
| **Marginal R-squared** | = | 0.252 | | | | |
| | Coef. | Std. Err. | t | P>\|t\| | [95% Conf. | Interval] |
| **Intervention** | -0.201 | 0.108 | -1.864 | 0.063 | -0.412 | 0.011 |
| **Daily Gas Use (kWh)** | -0.013 | 0.001 | -9.876 | <0.001 | -0.016 | -0.010 |
| **Daily Elec Use (kWh)** | -0.031 | 0.005 | -6.476 | <0.001 | -0.041 | -0.022 |
| **Floor size (log sqm)** | 1.234 | 0.403 | 3.061 | 0.002 | 0.442 | 2.026 |
| **2019 Group** | 0.319 | 0.173 | 1.838 | 0.067 | -0.022 | 0.659 |
| **2020 Group** | -0.254 | 0.206 | -1.236 | 0.217 | -0.659 | 0.150 |
| **Pre-winter star rating** | -0.153 | 0.031 | -4.894 | <0.001 | -0.215 | -0.092 |
| **Solar Panel (Y/N)** | 0.218 | 0.131 | 1.665 | 0.097 | -0.039 | 0.476 |
| **Intercept** | 0.556 | 0.892 | 0.624 | 0.533 | -1.196 | 2.309 |

*adjusted for LGA

Table 16 Linear regression for hours of Exposure to cold (ITT), overnight

| **Number of obs** | = | **662** | | | | |
|---|---|---|---|---|---|---|
| **Marginal R-squared** | = | 0.390 | | | | |
| | Coef. | Std. Err. | t | P>\|t\| | [95% Conf. | Interval] |
| **Intervention** | -0.275 | 0.173 | -0.614 | 0.064 | -1.594 | 0.112 |
| **Daily Gas Use (kWh)** | -0.033 | 0.002 | -0.037 | -0.028 | -14.342 | <0.001 |
| **Daily Elec Use (kWh)** | -0.108 | 0.009 | -0.125 | -0.091 | -12.281 | <0.001 |
| **Floor size (log sqm)** | 4.472 | 0.699 | 3.099 | 5.846 | 6.395 | <0.001 |
| **2019 Group** | 0.018 | 0.298 | -0.567 | 0.602 | 0.060 | 0.952 |
| **2020 Group** | 0.377 | 0.275 | -0.163 | 0.917 | 1.369 | 0.171 |
| **Pre-winter star rating** | -0.320 | 0.054 | -0.425 | -0.215 | -5.984 | <0.001 |
| **Solar Panel (Y/N)** | 0.535 | 0.226 | 0.092 | 0.979 | 2.369 | 0.018 |
| **Intercept** | 1.848 | 1.527 | -1.151 | 4.848 | 1.210 | 0.227 |

*adjusted for LGA

Table 17 Linear regression for hours of Exposure to cold (PP), overnight

| **Number of obs** | = | **512** |
|---|---|---|
| **Marginal R-squared** | = | 0.369 |

| | Coef. | Std. Err. | t | P>\|t\| | [95% Conf. | Interval] |
|---|---|---|---|---|---|---|
| **Intervention** | -0.234 | 0.204 | -1.150 | 0.251 | -0.634 | 0.166 |
| **Daily Gas Use (kWh)** | -0.032 | 0.003 | -12.603 | <0.001 | -0.037 | -0.027 |
| **Daily Elec Use (kWh)** | -0.111 | 0.009 | -12.059 | <0.001 | -0.130 | -0.093 |
| **Floor size (log sqm)** | 4.589 | 0.766 | 5.990 | <0.001 | 3.084 | 6.094 |
| **2019 Group** | -0.174 | 0.329 | -0.529 | 0.597 | -0.820 | 0.472 |
| **2020 Group** | 0.010 | 0.314 | 0.033 | 0.974 | -0.607 | 0.627 |
| **Pre-winter star rating** | -0.280 | 0.059 | -4.737 | <0.001 | -0.397 | -0.164 |
| **Solar Panel (Y/N)** | 0.417 | 0.249 | 1.672 | 0.095 | -0.073 | 0.907 |
| **Intercept** | 1.703 | 1.671 | 1.019 | 0.309 | -1.580 | 4.986 |

*adjusted for LGA

Table 18 Ordinal regression for perceived thermal comfort (ITT)

| **Number of obs** | = | **880** | | | | |
|---|---|---|---|---|---|---|
| **Marginal R-squared** | = | 0.539 | | | | |
| | Odds ratio. | Std. Err. | t | P>\|t\| | [95% Conf. | Interval] |
| **Intervention** | 2.344 | 0.299 | 6.689 | <0.001 | 1.826 | 3.009 |
| **2019 Group** | 1.060 | 0.248 | 0.249 | 0.803 | 0.671 | 1.675 |
| **2020 Group** | 1.339 | 0.380 | 1.028 | 0.304 | 0.768 | 2.336 |

*adjusted for LGA

Table 19 Linear regression for exposure to hours of hazardous conditions (ITT)

| **Number of obs** | = | **512** | | | | |
|---|---|---|---|---|---|---|
| **Marginal R-squared** | = | 0.264 | | | | |
| | Coef. | Std. Err. | t | P>\|t\| | [95% Conf. | Interval] |
| **Intervention** | -0.925 | 0.343 | -2.693 | 0.007 | -1.599 | -0.250 |
| **Daily Gas Use (kWh)** | -0.018 | 0.004 | -4.128 | <0.001 | -0.026 | -0.009 |
| **Daily Elec Use (kWh)** | -0.043 | 0.016 | -2.731 | 0.007 | -0.073 | -0.012 |
| **Floor size (log sqm)** | 1.716 | 1.292 | 1.328 | 0.185 | -0.823 | 4.254 |
| **2019 Group** | -5.505 | 0.555 | -9.920 | <0.001 | -6.595 | -4.414 |
| **2020 Group** | -6.574 | 0.530 | -12.408 | <0.001 | -7.615 | -5.533 |
| **Pre-winter star rating** | -0.146 | 0.100 | -1.458 | 0.146 | -0.342 | 0.051 |
| **Solar Panel (Y/N)** | 0.324 | 0.421 | 0.770 | 0.442 | -0.503 | 1.150 |
| **Intercept** | 6.591 | 2.818 | 2.339 | 0.020 | 1.054 | 12.127 |

*adjusted for LGA

Table 20 Linear regression for electricity use (ITT)

| **Number of obs** | = | **662** |
|---|---|---|
| **Marginal R-squared** | = | 0.192 |

| | Coef. | Std. Err. | t | P>\|t\| | [95% Conf. | Interval] |
|---|---|---|---|---|---|---|
| **Intervention** | -0.943 | 0.707 | -1.334 | 0.183 | -2.331 | 0.445 |
| **Indoor temperature** | 1.972 | 0.179 | 11.049 | <0.001 | 1.622 | 2.323 |
| **Daily Gas Use (kWh)** | -0.070 | 0.011 | -6.492 | <0.001 | -0.091 | -0.049 |
| **Floor size (log sqm)** | 17.055 | 2.854 | 5.977 | <0.001 | 11.452 | 22.658 |
| **2019 Group** | 1.487 | 1.215 | 1.224 | 0.221 | -0.898 | 3.871 |
| **2020 Group** | 1.853 | 1.121 | 1.653 | 0.099 | -0.349 | 4.054 |
| **Pre-winter star rating** | -0.714 | 0.225 | -3.178 | 0.002 | -1.154 | -0.273 |
| **Solar Panel (Y/N)** | 1.594 | 0.929 | 1.716 | 0.087 | -0.230 | 3.418 |
| **Intercept** | -52.760 | 7.138 | -7.391 | <0.001 | -66.776 | -38.743 |

*adjusted for LGA

Table 21 Linear regression for gas use (ITT)

| **Number of obs** | = **662** | | | | | |
|---|---|---|---|---|---|---|
| **Marginal R-squared** | = 0.437 | | | | | |
| | Coef. | Std. Err. | t | P>\|t\| | [95% Conf. | Interval] |
| **Intervention** | -7.080 | 2.485 | -2.849 | 0.005 | -11.959 | -2.201 |
| **Indoor temperature** | 9.568 | 0.576 | 16.605 | <0.001 | 8.437 | 10.699 |
| **Daily Elec Use (kWh)** | -0.870 | 0.134 | -6.487 | <0.001 | -1.133 | -0.606 |
| **Floor size (log sqm)** | 74.481 | 9.958 | 7.480 | <0.001 | 54.927 | 94.034 |
| **2019 Group** | -8.565 | 4.289 | -1.997 | 0.046 | -16.987 | -0.144 |
| **2020 Group** | -6.819 | 4.442 | -1.535 | 0.125 | -15.541 | 1.903 |
| **Pre-winter star rating** | -7.298 | 0.751 | -9.721 | <0.001 | -8.773 | -5.824 |
| **Solar Panel (Y/N)** | 14.926 | 3.244 | 4.601 | <0.001 | 8.556 | 21.295 |
| **Intercept** | -214.441 | 25.022 | -8.570 | <0.001 | -263.575 | -165.307 |

*adjusted for LGA

Table 22 Linear regression for MCS (ITT)

| **Number of obs** | = | **980** | | | | |
|---|---|---|---|---|---|---|
| **R-squared** | = | **0.3857** | | | | |
| | Coef. | Std. Err. | t | P>\|t\| | [95% Conf. | Interval] |
| **mcs0** | 0.515 | 0.028 | 18.210 | 0.000 | 0.460 | 0.571 |
| **Intervention** | 1.730 | 0.776 | 2.230 | 0.026* | 0.207 | 3.254 |
| **yr2019** | 1.692 | 1.446 | 1.170 | 0.242 | -1.146 | 4.530 |
| **yr2020** | 4.984 | 2.285 | 2.180 | 0.029* | 0.499 | 9.469 |
| **age_mncen** | 0.088 | 0.040 | 2.210 | 0.027* | 0.010 | 0.166 |
| **female** | -0.009 | 0.711 | -0.010 | 0.990 | -1.405 | 1.387 |
| **_cons** | 17.052 | 1.849 | 9.220 | 0.000 | 13.421 | 20.682 |

Notes: Age is mean centred. The baseline data (**mcs0**) is controlled for the self-reported outcomes. LGAs are included in the regressions which are not reported in the tables. +p<0.10, *p<0.05, **p<.0.01, ***p<0.001.

Table 23 Linear regression for PCS (ITT)

| **Number of obs** | = | **980** | | | | |
|---|---|---|---|---|---|---|
| **R-squared** | = | **0.4531** | | | | |
| | Coef. | Std. Err. | t | P>\|t\| | [95% Conf. | Interval] |
| **pcs0** | 0.714 | 0.029 | 24.220 | 0.000 | 0.656 | 0.772 |
| **Intervention** | 0.808 | 0.565 | 1.430 | 0.153 | -0.300 | 1.916 |
| **2019** | 3.061 | 0.885 | 3.460 | 0.001* | 1.324 | 4.798 |
| **2020** | 0.254 | 1.674 | 0.150 | 0.880 | -3.032 | 3.539 |
| **age_mncen** | -0.064 | 0.028 | -2.250 | 0.025* | -0.119 | -0.008 |
| **female** | -0.978 | 0.605 | -1.620 | 0.106 | -2.165 | 0.208 |
| **_cons** | 8.634 | 1.297 | 6.660 | 0.000 | 6.088 | 11.179 |

Notes: Age is mean centred. The baseline data (**pcs0**) is controlled for the self-reported outcomes. LGAs are included in the regressions which are not reported in the tables. +p<0.10, *p<0.05, **p<.0.01, ***p<0.001.

Table 24: Linear regression results for ASCOT score (ITT)

| **Number of obs** | = | **1,072** | | | | |
|---|---|---|---|---|---|---|
| **R-squared** | = | **0.3765** | | | | |
| | Coef. | Std. Err. | t | P>\|t\| | [95% Conf. | Interval] |
| **ascot_ut0** | 0.575 | 0.034 | 16.980 | 0.000 | 0.509 | 0.642 |
| **Intervention** | 0.024 | 0.009 | 2.640 | 0.009* | 0.006 | 0.042 |
| **yr2019** | 0.006 | 0.016 | 0.350 | 0.725 | -0.026 | 0.037 |
| **yr2020** | 0.014 | 0.024 | 0.610 | 0.543 | -0.032 | 0.061 |
| **age_mncen** | 0.001 | 0.000 | 1.260 | 0.208 | 0.000 | 0.001 |
| **female** | 0.007 | 0.008 | 0.870 | 0.386 | -0.009 | 0.024 |
| **_cons** | 0.316 | 0.031 | 10.240 | 0.000 | 0.255 | 0.376 |

Notes: Age is mean centred. The baseline data (ascot_ut0) is controlled for the self-reported outcomes. LGAs are included in the regressions which are not reported in the tables. +p<0.10, *p<0.05, **p<.0.01, ***p<0.001.

Table 25 Linear regression results for predicting EQ-5D-5L scores (ITT)

| **Number of obs** | = | **1,110** | | | | |
|---|---|---|---|---|---|---|
| **R-squared** | = | **0.3536** | | | | |
| | Coef. | Std. Err. | t | P>\|t\| | 95% CI | |
| **eq5d5lscore0** | 0.553 | 0.029 | 19.050 | 0.000 | 0.496 | 0.610 |
| **Intervention** | 0.009 | 0.017 | 0.530 | 0.597 | -0.025 | 0.043 |
| **yr2019** | 0.047 | 0.032 | 1.440 | 0.150 | -0.017 | 0.110 |
| **yr2020** | 0.091 | 0.044 | 2.060 | 0.040* | 0.004 | 0.178 |
| **age_mncen** | -0.001 | 0.001 | -1.570 | 0.117 | -0.003 | 0.000 |
| **Female** | 0.006 | 0.018 | 0.330 | 0.742 | -0.030 | 0.041 |
| **_cons** | 0.185 | 0.036 | 5.190 | 0.000 | 0.115 | 0.255 |

Notes: Age is mean centred. The baseline data (eq5d5lscore0) is controlled for the self-reported outcomes. LGAs are included in the regressions which are not reported in the tables. +p<0.10, *p<0.05, **p<.0.01, ***p<0.001.

Table 26: Linear regression outputs for 'health today' scores (ITT)

| **Number of obs** | = | **1,091** | | | | |
|---|---|---|---|---|---|---|
| **R-squared** | = | **0.2755** | | | | |
| | Coef. | Std. Err. | t | P>t | [95% Conf. | Interval] |
| **healthtoday0** | 0.516 | 0.031 | 16.630 | 0.000 | 0.455 | 0.577 |
| **intervention** | 1.063 | 1.193 | 0.890 | 0.373 | -1.278 | 3.404 |
| **yr2019** | 0.736 | 2.133 | 0.340 | 0.730 | -3.451 | 4.923 |
| **yr2020** | 5.313 | 2.899 | 1.830 | 0.067+ | -0.377 | 11.002 |
| **age** | 0.038 | 0.053 | 0.720 | 0.473 | -0.065 | 0.141 |
| **female** | -1.007 | 1.142 | -0.880 | 0.378 | -3.248 | 1.235 |
| **_cons** | 27.349 | 4.853 | 5.640 | 0.000 | 17.825 | 36.873 |

Notes: Age is mean centred. The baseline data (healthtoday0) is controlled for the self-reported outcomes. LGAs are included in the regressions which are not reported in the tables. +p<0.10, *p<0.05, **p<.0.01, ***p<0.001.

Table 27: Negative binomial regression results for predicting MBS services (ITT)

| **Negative binomial regression** | | | | **Number of obs =** | | **1,238** |
|---|---|---|---|---|---|---|
| | | | | **Wald chi2(13) =** | | **19.06** |
| **Dispersion = mean** | | | | **Prob > chi2 =** | | **0.1213** |
| **Log pseudolikelihood = -4412.8709** | | | | **Pseudo R2 =** | | **0.0035** |
| **MBSservice** | **IRR** | Std. Err. | z | P>z | [95% Conf. | Interval] |
| **group2** | 0.906841 | 0.050551 | -1.75 | 0.079 | 0.812983 | 1.011533 |
| **age** | 1.003179 | 0.002673 | 1.19 | 0.233 | 0.997955 | 1.008432 |
| **sex** | 0.992382 | 0.051042 | -0.15 | 0.882 | 0.897219 | 1.097638 |
| **yr2019** | 1.130875 | 0.108226 | 1.29 | 0.199 | 0.937463 | 1.364192 |
| **yr2020** | 0.855486 | 0.105526 | -1.27 | 0.206 | 0.671763 | 1.089457 |
| **_cons** | 11.52773 | 2.522531 | 11.17 | 0 | 7.507261 | 17.70134 |
| **/lnalpha** | -0.55327 | 0.068603 | | | -0.68773 | -0.41881 |
| **alpha** | 0.575068 | 0.039451 | | | 0.502718 | 0.657831 |

Table 28: Negative binomial regression results for predicting PBS services (ITT)

| **Number of obs = 1,162** | | | | | | |
|---|---|---|---|---|---|---|
| **Pseudo R2 = 0.0024** | | | | | | |
| **Std. Err. adjusted for 907 clusters in hhid** | | | | | | |
| **PBSservice** | Coef. | Std. Err. | z | P>z | 95% CI LB | 95% CI UB |
| **Intervention** | 0.019 | 0.038 | 0.490 | 0.622 | -0.055 | 0.093 |
| **yr2019** | -0.057 | 0.071 | -0.800 | 0.423 | -0.196 | 0.082 |
| **yr2020** | -0.097 | 0.108 | -0.900 | 0.370 | -0.309 | 0.115 |

| | | | | | | |
|---|---|---|---|---|---|---|
| **age** | 0.004 | 0.002 | 1.740 | 0.082+ | 0.000 | 0.008 |
| **sex** | -0.083 | 0.037 | -2.260 | 0.024* | -0.156 | -0.011 |
| **_cons** | 2.566 | 0.178 | 14.410 | 0.000 | 2.217 | 2.916 |
| **/lnalpha** | -1.133 | 0.053 | | | -1.238 | -1.028 |
| **alpha** | 0.322 | 0.017 | | | 0.290 | 0.358 |

Notes: Notes: LGAs are included in the regressions which are not reported in the tables. +p<0.10, *p<0.05, **p<.0.01, ***p<0.001. *The dispersion parameter, **alpha**, is significantly greater than zero than the data are over dispersed and are better estimated using a negative binomial model than a poisson model.*

Table 29: Negative binomial regression results for predicting GP services (ITT)

| **Number of obs=1,238** | | | | | | |
|---|---|---|---|---|---|---|
| **Pseudo R2= 0.0189** | | | | | | |
| **Std. Err. adjusted for 952 clusters in hhid** | | | | | | |
| | Coef. | Std. Err. | z | P>z | 95% CI LB | 95% CI UB |
| **Intervention** | 0.017 | 0.064 | 0.260 | 0.793 | -0.109 | 0.142 |
| **yr2019** | 0.026 | 0.093 | 0.280 | 0.779 | -0.156 | 0.208 |
| **yr2020** | -0.494 | 0.253 | -1.950 | 0.051+ | -0.990 | 0.001 |
| **age** | 0.003 | 0.003 | 1.180 | 0.239 | -0.002 | 0.009 |
| **sex** | -0.071 | 0.064 | -1.120 | 0.265 | -0.196 | 0.054 |
| **_cons** | 1.052 | 0.238 | 4.420 | 0.000 | 0.585 | 1.519 |
| **/lnalpha** | -0.452 | 0.084 | | | -0.617 | -0.287 |
| **alpha^** | 0.636 | 0.054 | | | 0.540 | 0.751 |

Notes: LGAs are included in the regressions which are not reported in the tables. +p<0.10, *p<0.05, **p<.0.01, ***p<0.001. *The dispersion parameter, **alpha**, is significantly greater than zero than the data are over dispersed and are better estimated using a negative binomial model than a poisson model.*

Table 30: Linear regression results for predicting MBS charges (ITT)

| **MBS charge** | Coef. | Std. Err. | t | P>t | [95% Conf. | Interval] |
|---|---|---|---|---|---|---|
| **Intervention** | -156.573 | 78.525 | -1.990 | 0.046* | -310.675 | -2.471 |
| **yr2019** | 197.508 | 135.796 | 1.450 | 0.146 | -68.988 | 464.003 |
| **yr2020** | -280.853 | 114.515 | -2.450 | 0.014* | -505.584 | -56.121 |
| **age** | 1.174 | 3.605 | 0.330 | 0.745 | -5.901 | 8.249 |
| **sex** | 59.936 | 65.749 | 0.910 | 0.362 | -69.094 | 188.966 |
| **_cons** | 818.951 | 300.540 | 2.720 | 0.007 | 229.152 | 1408.750 |

Notes: LGAs are included in the regressions which are not reported in the tables. +p<0.10, *p<0.05, **p<.0.01, ***p<0.001.

Table 31: Linear regression results for predicting MBS benefits (ITT)

| | | | | | | |
|---|---|---|---|---|---|---|
| **Intervention** | -107.871 | 62.318 | -1.730 | 0.084+ | -230.168 | 14.426 |
| **yr2019** | 85.087 | 101.137 | 0.840 | 0.400 | -113.391 | 283.566 |

| | | | | | | |
|---|---|---|---|---|---|---|
| **yr2020** | -235.939 | 96.570 | -2.440 | 0.015* | -425.454 | -46.424 |
| **age** | 1.942 | 2.331 | 0.830 | 0.405 | -2.633 | 6.517 |
| **sex** | 18.132 | 53.734 | 0.340 | 0.736 | -87.319 | 123.583 |
| **_cons** | 713.409 | 196.953 | 3.620 | 0.000 | 326.896 | 1099.921 |

Notes: LGAs are included in the regressions which are not reported in the tables. +p<0.10, *p<0.05, **p<.0.01, ***p<0.001.

Table 30: Linear regression results for predicting MBS out-of-pocket costs (ITT)

| **Out-of-pocket** | **Coef.** | **Std. Err.** | **T** | **P>t** | **95% CI LB** | **95% CI UB** |
|---|---|---|---|---|---|---|
| **Intervention** | -48.702 | 24.932 | -1.950 | 0.051+ | -97.629 | 0.225 |
| **yr2019** | 112.421 | 47.546 | 2.360 | 0.018* | 19.112 | 205.729 |
| **yr2020** | -44.914 | 32.894 | -1.370 | 0.172 | -109.467 | 19.638 |
| **age** | -0.768 | 1.653 | -0.460 | 0.642 | -4.012 | 2.475 |
| **sex** | 41.804 | 19.148 | 2.180 | 0.029* | 4.227 | 79.381 |
| **_cons** | 105.543 | 135.472 | 0.780 | 0.436 | -160.316 | 371.401 |

Notes: LGAs are included in the regressions which are not reported in the tables. +p<0.10, *p<0.05, **p<.0.01, ***p<0.001.

Table 32: Linear regression results for predicting total gross price for PBS (ITT)

| **F(13, 906) = 1.29** | | | | | | |
|---|---|---|---|---|---|---|
| **Prob > F = 0.2116** | | | | | | |
| **R-squared = 0.0201** | | | | | | |
| **Root MSE = 1687.7** | | | | | | |
| **gross_price** | Coef. | Std. Err. | t | P>t | 95% CI LB | 95% CI UB |
| **Intervention** | -70.940 | 108.037 | -0.660 | 0.512 | -282.971 | 141.091 |
| **yr2019** | 7.459 | 108.262 | 0.070 | 0.945 | -205.013 | 219.932 |
| **yr2020** | -94.761 | 122.652 | -0.770 | 0.440 | -335.476 | 145.953 |
| **age** | -1.052 | 5.186 | -0.200 | 0.839 | -11.230 | 9.125 |
| **sex** | -245.267 | 121.546 | -2.020 | 0.044 | -483.811 | -6.723 |
| **_cons** | 872.805 | 360.907 | 2.420 | 0.016 | 164.494 | 1581.116 |

Notes: LGAs are included in the regressions which are not reported in the tables. +p<0.10, *p<0.05, **p<.0.01, ***p<0.001.

Table 33: Linear regression results for predicting net benefits paid

| **Number of obs = 1,162** | | | | | | |
|---|---|---|---|---|---|---|
| **F(13, 906) = 1.21** | | | | | | |
| **Prob > F = 0.2655** | | | | | | |
| **R-squared = 0.021** | | | | | | |
| **Root MSE = 1684.8** | | | | | | |
| **net_ben** | Coef. | Std. Err. | t | P>t | 95% CI LB | 95% CI UB |
| **Intervention** | -71.360 | 108.079 | -0.660 | 0.509 | -283.473 | 140.754 |
| **yr2019** | 12.221 | 108.640 | 0.110 | 0.910 | -200.994 | 225.436 |
| **yr2020** | -69.104 | 123.692 | -0.560 | 0.577 | -311.860 | 173.653 |
| **age** | -1.174 | 5.180 | -0.230 | 0.821 | -11.341 | 8.992 |
| **sex** | -255.540 | 121.298 | -2.110 | 0.035* | -493.597 | -17.484 |
| **_cons** | 817.879 | 360.126 | 2.270 | 0.023 | 111.101 | 1524.657 |

Notes: LGAs are included in the regressions which are not reported in the tables. +p<0.10, *p<0.05, **p<.0.01, ***p<0.001.

Table 35: Linear regression results for predicting patient contribution for PBS (ITT)

| | | | | | | |
|---|---|---|---|---|---|---|
| **Number of obs = 1,162** | | | | | | |
| **F(13, 906) = 3.29** | | | | | | |
| **Prob > F = 0.0001** | | | | | | |
| **R-squared = 0.0385** | | | | | | |
| **Root MSE = 51.179** | | | | | | |
| **patient_cont** | Coef. | Std. Err. | t | P>t | 95% CI LB | 95% CI UB |
| **Intervention** | 0.420 | 3.267 | 0.130 | 0.898 | -5.993 | 6.832 |
| **yr2019** | -4.762 | 5.489 | -0.870 | 0.386 | -15.534 | 6.010 |
| **yr2020** | -25.658 | 7.903 | -3.250 | 0.001** | -41.168 | -10.147 |
| **age** | 0.122 | 0.154 | 0.800 | 0.426 | -0.179 | 0.424 |
| **sex** | 10.273 | 3.053 | 3.370 | 0.001** | 4.282 | 16.265 |
| **_cons** | 54.926 | 12.402 | 4.430 | 0.000 | 30.586 | 79.266 |

Notes: LGAs are included in the regressions which are not reported in the tables. +p<0.10, *p<0.05, **p<.0.01, ***p<0.001.

Table 36: Regression predicting the total number of hospital admissions (ITT)

| | | | | | | |
|---|---|---|---|---|---|---|
| **Negative binomial regression** | | | | **Number of obs** | | **= 1,295** |
| **Dispersion = mean** | | | | **Wald chi2(13)** | | **= 27.42** |
| **Log pseudolikelihood = -1016.2568** | | | | **Prob > chi2** | | **= 0.0109** |
| **Adjusted for 985 cluster in hhid** | | | | **Pseudo R2** | | **= 0.0273** |
| **Hospitalisations** | Coef. | Std. Err. | z | P>z | [95% Conf. | Interval] |
| **Intervention** | -0.172 | 0.205 | -0.84 | 0.401 | -0.574 | 0.229 |
| **year2019** | 0.367 | 0.298 | 1.23 | 0.218 | -0.217 | 0.951 |
| **year2020** | 0.149 | 0.598 | 0.25 | 0.803 | -1.024 | 1.322 |
| **age** | -0.003 | 0.008 | -0.36 | 0.718 | -0.020 | 0.013 |
| **sex** | -0.754 | 0.222 | -3.40 | 0.001 | -1.188 | -0.319 |
| **_cons** | -0.383 | 0.707 | -0.54 | 0.588 | -1.769 | 1.002 |
| **/lnalpha** | 1.926 | 0.121 | | 1.688 | 2.164 | |
| **alpha** | 6.862 | 0.833 | | 5.409 | 8.706 | |

Notes: LGAs are included in the regressions which are not reported in the tables. +p<0.10, *p<0.05, **p<.0.01, ***p<0.001. *The dispersion parameter, **alpha**, is significantly greater than zero than the data are over dispersed and are better estimated using a negative binomial model than a poisson model.*

Table 37: Regression results for outcome of hospital cost (ITT)

| | | | | | | |
|---|---|---|---|---|---|---|
| **Number of obs =1,295** | | | | | | |
| **R-squared =** | | **0.0130** | | | | |
| **Hospital Cost** | Coef. | Std. Err. | t | P>t | [95% Conf. | Interval] |

| | | | | | | |
|---|---|---|---|---|---|---|
| **Intervention** | -557.477 | 438.199 | -1.27 | 0.204 | -1417.399 | 302.444 |
| **year2019** | 158.637 | 748.950 | 0.21 | 0.832 | -1311.103 | 1628.377 |
| **year2020** | -1265.974 | 956.792 | -1.32 | 0.186 | -3143.583 | 611.635 |
| **age** | 2.429 | 18.497 | 0.13 | 0.896 | -33.869 | 38.727 |
| **sex** | -1097.395 | 503.969 | -2.18 | 0.030 | -2086.385 | -108.406 |
| **_cons** | 3149.012 | 1587.758 | 1.98 | 0.048 | 33.195 | 6264.828 |

Notes: LGAs are included in the regressions which are not reported in the tables. +p<0.10, *p<0.05, **p<.0.01, ***p<0.001.

Table 38: Regression results for outcome of length of stay (ITT)

| **Negative binomial regression** | | | | | **Number of obs =** | **1,295** |
|---|---|---|---|---|---|---|
| | | | | | **Wald chi2(13) =** | **13.78** |
| **Dispersion = mean** | | | | | **Prob > chi2 =** | **0.3893** |
| **Log pseudolikelihood = -962.55096** | | | | | **Pseudo R2 =** | **0.0055** |
| **Std. Err. adjusted for 976 clusters in hhid** | | | | | | |
| **Length of Stay** | Coef. | Std. Err. | z | P>z | [95% Conf. | Interval] |
| **Intervention** | 0.123 | 0.233 | 0.53 | 0.597 | -0.334 | 0.580 |
| **year2019** | -0.340 | 0.380 | -0.89 | 0.371 | -1.086 | 0.405 |
| **year2020** | -1.407 | 0.711 | -1.98 | 0.048 | -2.800 | -0.014 |
| **age** | 0.002 | 0.012 | 0.20 | 0.840 | -0.022 | 0.027 |
| **sex** | -0.321 | 0.268 | -1.20 | 0.231 | -0.847 | 0.204 |
| **_cons** | 0.552 | 1.035 | 0.53 | 0.594 | -1.476 | 2.579 |
| **/lnalpha** | 3.203 | 0.094 | | 3.019 | 3.387 | |
| **alpha** | 24.613 | 2.312 | | 20.474 | 29.588 | |

Notes: LGAs are included in the regressions which are not reported in the tables. +p<0.10, *p<0.05, **p<.0.01, ***p<0.001. *The dispersion parameter, **alpha**, is significantly greater than zero than the data are over dispersed and are better estimated using a negative binomial model than a poisson model.*

Table 39: Regression model predicting total number of ED visits (ITT)

| **Negative binomial regression** | | | **Number of obs** | | | **= 1,293** |
|---|---|---|---|---|---|---|
| | | | **Wald chi2(13)** | | | **= 25.77** |
| **Dispersion = mean** | | | **Prob > chi2** | | | **= 0.0182** |
| **Log pseudolikelihood = -699.73343** | | | **Pseudo R2** | | | **= 0.0172** |
| | | | **(Std. Err. adjusted for 975 clusters in hhid)** | | | |
| **Admissions (total)** | Coef. | Std. Err. | z | P>z | [95% Conf. | Interval] |
| **Intervention** | 0.030 | 0.171 | 0.18 | 0.861 | -0.305 | 0.364 |
| **year2019** | -0.566 | 0.287 | -1.97 | 0.049 | -1.129 | -0.002 |
| **year2020** | -0.953 | 0.429 | -2.22 | 0.026 | -1.795 | -0.112 |
| **age** | -0.007 | 0.008 | -0.97 | 0.332 | -0.022 | 0.008 |
| **sex** | -0.423 | 0.168 | -2.52 | 0.012 | -0.751 | -0.094 |

| | | | | | | |
|---|---|---|---|---|---|---|
| **_cons** | -0.375 | 0.627 | -0.60 | 0.550 | -1.604 | 0.855 |
| **/lnalpha** | 1.511 | 0.151 | | 1.215 | 1.807 | |
| **alpha** | 4.531 | 0.685 | | 3.370 | 6.092 | |

Notes: LGAs are included in the regressions which are not reported in the tables. +p<0.10, *p<0.05, **p<.0.01, ***p<0.001. *The dispersion parameter, **alpha**, is significantly greater than zero than the data are over dispersed and are better estimated using a negative binomial model than a poisson model.*

Table 40: Regression results for outcome of ED cost (ITT)

| **Number of obs =** | **1,293** | | | | | |
|---|---|---|---|---|---|---|
| **R-squared =** | **0.0223** | | | | | |
| **Cost** | Coef. | Std. Err. | t | P>t | [95% Conf. | Interval] |
| **Intervention** | -4.199 | 16.197 | -0.26 | 0.796 | -35.984 | 27.586 |
| **year2019** | -68.945 | 36.095 | -1.91 | 0.056 | -139.779 | 1.888 |
| **year2020** | -40.105 | 45.849 | -0.87 | 0.382 | -130.080 | 49.870 |
| **age** | -0.743 | 0.805 | -0.92 | 0.356 | -2.322 | 0.836 |
| **sex** | -54.129 | 18.669 | -2.90 | 0.004 | -90.765 | -17.494 |
| **_cons** | 208.274 | 69.822 | 2.98 | 0.003 | 71.256 | 345.293 |

Notes: LGAs are included in the regressions which are not reported in the tables. +p<0.10, *p<0.05, **p<.0.01, ***p<0.001.

Table 41: Regression predicting total service use for (ITT)

| **Total Service** | Coef. | Std. Err. | t | P>t | [95% Conf. | Interval] |
|---|---|---|---|---|---|---|
| **Intervention** | -1.29 | 1.21 | -1.07 | 0.29 | -3.67 | 1.08 |
| **age** | 0.13 | 0.05 | 2.44 | 0.02* | 0.03 | 0.23 |
| **sex** | -1.49 | 1.16 | -1.29 | 0.20 | -3.77 | 0.78 |
| **yr2019** | 1.00 | 2.14 | 0.47 | 0.64 | -3.19 | 5.20 |
| **yr2020** | -3.33 | 2.67 | -1.25 | 0.21 | -8.57 | 1.91 |
| **_cons** | 21.09 | 4.50 | 4.69 | 0.00 | 12.27 | 29.92 |

Notes: LGAs are included in the regressions which are not reported in the tables. +p<0.10, *p<0.05, **p<.0.01, ***p<0.001. *T*

Table 42: Regression predicting total service costs for (ITT)

| **totalcosts** | Coef. | Std. Err. | t | P>t | [95% Conf. | Interval] |
|---|---|---|---|---|---|---|
| **Intervention** | -886.51 | 505.87 | -1.75 | 0.08+ | -1879.25 | 106.23 |
| **age** | -0.19 | 22.36 | -0.01 | 0.99 | -44.07 | 43.70 |
| **sex** | -1315.57 | 570.85 | -2.30 | 0.02 | -2435.84 | -195.30 |
| **yr2019** | 306.53 | 849.82 | 0.36 | 0.72 | -1361.20 | 1974.26 |
| **yr2020** | -1264.71 | 1143.82 | -1.11 | 0.27 | -3509.40 | 979.98 |
| **_cons** | 5166.14 | 1891.83 | 2.73 | 0.01 | 1453.54 | 8878.74 |

Notes: LGAs are included in the regressions which are not reported in the tables. +p<0.10, *p<0.05, **p<.0.01, ***p<0.001.

Table 42: Regression predicting absenteeism (from usual activities) for (ITT)

| **absenteeism** | Coef. | IRR | Std. Err. | z | P>z | 95% CI | |
|---|---|---|---|---|---|---|---|
| **Intervention** | -0.22 | 0.80 | 0.21 | -1.08 | 0.28 | -0.62 | 0.18 |
| **yr2019** | -0.38 | 0.68 | 0.36 | -1.06 | 0.29 | -1.09 | 0.32 |
| **yr2020** | -1.05 | 0.35 | 0.55 | -1.92 | $0.06^{+}$ | -2.12 | 0.02 |
| **_cons** | 2.69 | 14.77 | 0.31 | 8.60 | 0.00 | 2.08 | 3.31 |
| **/lnalpha** | 2.21 | 2.21 | 0.07 | | | 2.08 | 2.35 |
| **alpha** | 9.13 | 9.13 | 0.62 | | | 7.99 | 10.44 |

*age and sex not accounted for in model because it was a household level measure (all eligible householders)

Table 43: Regression predicting respiratory symptoms for (ITT)

| **Ordered logistic regression Number of obs = 1,046** | | | | | | |
|---|---|---|---|---|---|---|
| **Wald chi2(13) = 41.46** | | | | | | |
| **Prob > chi2 = 0.0001** | | | | | | |
| **Log pseudolikelihood = -1603.3861 Pseudo R2 = 0.0117** | | | | | | |
| **(Std. Err. adjusted for 838 clusters in HouseholdID)** | | | | | | |
| | | Robust | | | | |
| **mrc_difference** | Coef. | Std. Err. | z | P>z | [95% Conf. | Interval] |
| | -0.379 | 0.116 | -3.260 | 0.001** | -0.607 | -0.152 |
| **Intervention** | -0.321 | 0.208 | -1.550 | 0.122 | -0.728 | 0.086 |
| **yr2019** | 0.207 | 0.281 | 0.740 | 0.460 | -0.343 | 0.758 |
| **yr2020** | 0.012 | 0.005 | 2.250 | 0.024* | 0.002 | 0.022 |
| **age_mncen** | 0.066 | 0.119 | 0.550 | 0.582 | -0.168 | 0.300 |
| **female** | 0.075 | 0.134 | 0.560 | 0.576 | -0.188 | 0.339 |
| **/cut1** | -4.737 | 0.674 | | | -6.057 | -3.416 |
| **/cut2** | -3.236 | 0.502 | | | -4.220 | -2.251 |
| **/cut3** | -1.787 | 0.446 | | | -2.662 | -0.913 |
| **/cut4** | -0.413 | 0.435 | | | -1.267 | 0.440 |
| **/cut5** | 1.536 | 0.442 | | | 0.670 | 2.402 |
| **/cut6** | 2.852 | 0.453 | | | 1.963 | 3.740 |
| **/cut7** | 4.175 | 0.485 | | | 3.225 | 5.125 |
